# Estimates of the ongoing need for social distancing and control measures post-“lockdown” from trajectories of COVID-19 cases and mortality

**DOI:** 10.1101/2020.04.26.20080994

**Authors:** Mike Lonergan, James Chalmers

## Abstract

By 29^th^ April 2020, COVID-19 had caused more than 3 million cases across more than 200 countries. And most countries with significant outbreaks had introduced social distancing or “lockdown” measures to reduce viral transmission. So the key question now is when, how, and to what extent, these measures can be lifted.

By fitting regression models to publically available data on daily numbers of newly-confirmed cases and mortality, trajectories, doubling times and reproduction number (R_0_) were estimated both before and under the control measures. These data ran up to 29th April 2020, and covered 73 countries that had provided sufficient data for modelling.

The estimates of R_0_, before lockdown, based on these data were broadly consistent with those previously published at between 2.0 and 3.7 in the countries with the largest number of cases available for analysis (USA, Italy, Spain, France and UK). There was little evidence to suggest that the restrictions had reduced R far below 1 in many places, with France having the most rapid reductions – R_0_ 0.77 (95%CI 0.68-0.87), based on cases and 0.78 (95%CI 0.68-0.88) based on mortality.

Intermittent lockdown has been proposed as a means of controlling the outbreak while allowing periods of increase freedom and economic activity. These data suggest that few countries could have even one week per month unrestricted without seeing resurgence of the epidemic. Similarly, restoring 20% of the activity that has been prevented by the lockdowns looks difficult to reconcile with preventing the resurgence of the disease in most countries.

## Introduction

COVID-19 is a respiratory disease caused by a novel coronavirus (SARS-CoV-2).^1^ The spread of COVID-19 has already been the defining event of 2020.^1–3^ Over the past two decades, two other emerging pathogenic coronaviruses capable of causing life-threatening disease in humans and animals had been identified, namely severe acute respiratory syndrome coronavirus (SARS-CoV) and Middle Eastern respiratory syndrome coronavirus (MERS-CoV).^4^ While both of these prior novel coronaviruses caused significant outbreaks, they appear to have been more lethal and less transmissible and did not cause pandemics.

SARS-CoV-2 in contrast has spread through most of the world and caused most countries to restrict travel, close large parts of industry, restructure economies, and focus on disease control. Up to 29^th^ April there have been 3,053,457 cases of COVID-19 confirmed and 214,862 deaths in individuals testing positive.^5^ Cases have been reported in 207 countries.

“Lockdown” measures required to control the spread of the virus have been expensive in terms of reducing economic activity and painful through restricting social interaction, but, in late-April, there continue to be large numbers of mortalities in most parts of the world.^5^ The question of when and how far to ease the restrictions is, therefore, urgent. Planning how and when to lift restrictions requires an understanding of the transmissibility of the virus and effectiveness of the social distancing and lockdown measures taken across the world to date. The difference between the spread of the virus without restrictions and the spread or reduction in rates of infection under lockdown allow an estimate of how effective lockdown measures have been, and therefore the degree to which these measures can be relaxed without disease resurgence.

This paper uses publically available data from multiple countries to model the spread of COVID-19, both before and under the lockdown, and estimate the scope for relaxing the current restrictions.

## Methods

### Data

Data on the numbers of new confirmed cases of COVID-19, and numbers of deaths reported for people known to have COVID-19, for each day up to 29^th^ April 2020, were obtained from the European Centre for Disease Prevention and Control website (https://www.ecdc.europa.eu/en/publications-data/download-todays-data-geographic-distribution-COVID-19-19-cases-worldwide) on the 29^th^ April. Mortality data was used for countries that had reported at least 100 deaths, and numbers of cases for those where at least 1000 confirmed cases had been reported. These data series were trimmed, discarding days before a total of 10 deaths or 100 cases had been reported. That left 73 countries with sufficient data to fit models.

Both these types of data have important limitations, missing varying proportions of individuals in each country and over time. The limitations and changes in testing strategies make the data on confirmed cases particularly problematic, while the mortality data have an intrinsic lag that reduces their sensitivity to recent changes. Nevertheless, most countries have systematic testing of hospitalized cases and most mortality occurs in hospitals, making disease specific mortality potentially less biased by testing policies that confirmed cases. The mortality data is therefore prioritised in the interpretation when the results conflict. Data for each included country were carefully reviewed for inconsistencies and artefacts. The data for China on 18th April showed a spike that appears to be an artefact of redefinition, and was discarded. Iran reported nothing on the 4^th^ April and a spike on the 5^th^, so those reports were split evenly across the two days.

### Analysis

The data were loaded into R 3.6.1 (R Core Team 2019)^6^ and separate sets of regression models were fitted for each of mortality and cases in each of the countries. Preliminary examination of the data showed that residuals around models fitted to almost all the datasets were overdispersed relative to the Poisson distribution. All the models therefore used the negative binomial error family, with a log link function.

The aim was to calculate exponential growth rates for each country before and under lockdown. Each country introduced restrictions in a different pattern, so rather than attempting to interpret those rules, and how people responded to them, the data were used to identify periods of steady increase and decline. While the absolute numbers of cases and deaths are shocking, the daily numbers are very small proportions of the national populations and therefore the numbers of susceptible individuals are almost constant. This means that during periods of constant behaviour exponential trajectories can be expected, whether these trajectories are increasing or decreasing.

Two generalised linear models were fitted for each combination of start and finish date at least 10 days apart. The first model simply contained a linear term for time, the second one also had a quadratic term for time. BIC, the Bayesian information criterion^7^ was calculated and compared for each pair of models. The choice of BIC as the criterion for identifying exponential periods was entirely pragmatic. Exploratory analyses using Akiake’s Information Criterion (AIC)^8^ showed this selected very short intervals, and the small sample correction, AICc, worsens this problem. In BIC the penalty for additional parameters is proportional to the natural logarithm of the number of datapoints, so it is less eager to increase the complexity of models as sample size increases.

Linear models were considered potential representations of the initial exponential growth phase if they: i) used data ending no later than 5 days before the day that the maximum number of deaths, or cases, was observed; 2) had positive point estimates of slope; and 3) had BIC lower than their parallel quadratic model. If multiple models satisfied these criteria, the model where BIC was furthest below that of its quadratic equivalent was used. The model for the exponential period under lockdown was similarly chosen, but with the requirements that its data not begin before five days after the finish of that for the first model and its BIC be lower than that of its quadratic equivalent. No requirement was placed on its slope, so this could be negative or positive, and even larger than that for the first period, if the data suggested that the spread of the disease had accelerated. One exponential period was identified in the case data from 18 countries, and two exponential periods for 53 others. For the mortality data, these numbers were 3 and 35.

The five day interval was chosen after exploration of the Spanish and Italian data, where tight lockdowns were associated with obvious changes in the trajectory. These changes were particularly visible because of the large numbers of infections in those countries at that time. This interval does not need capture the whole course of multiple generations of infections, it merely needs to contain the bulk of the time over which the age-structure, in days since catching, of the infected individuals changes. Adjusting this interval changes the estimates, and estimability, of individual countries’ trajectories but produces similar overall patterns.

To provide a visual check on these models, a generalised additive model (GAM) of the whole trajectory was also fitted^9^. In most cases, the ends of this curve are similar to the exponential pattern match, and it is not surprising that the transition period does not. It should be noted that GAMs favour steady patterns of change and curvature, while many of the changes in behaviour were quite sudden. For a few countries the mismatch between the GAM and exponential models gave, subjectively, cause for concern. These are indicated in the figures.

The slopes, and confidence intervals around them, are of limited direct use. However, dividing the natural logarithm of 2 by them gives the doubling time of epidemic growth or halving time of its decay.

### Reproduction (R) number and estimates of effect of “lockdown”

R_0_, the basic reproduction number for the disease, is the expected number of people one infected person would cause in a naïve population. It is critical to disease spread: if it is above 1 an epidemic will accelerate, below that the outbreak will fade away. Because these data include only a fraction of the cases, R_0_ in each country, cannot be directly estimated from them. Instead, the R0 library^10^ was used to apply the method of Wallinga and Lipsitch^11^ to convert the estimates, and associated uncertainties, of the exponential trajectories into estimates of R_0_ both before and under lockdown. This approach requires an estimated distribution for the serial interval of infection. The lognormal with mean 4 days and standard deviation of 2.9 days calculated by Nishiura, Linton and Akhmetzhanov^12^ was used. While changing this distribution changes the individual estimated values for R_0_, the relationship between the estimates before and under lockdown is relatively insensitive to plausible choices.

Three estimates of the scope for relaxing lockdown were then considered. The first was a simple ratio of the slopes before and under lockdown. This indicates the number of days under lockdown required to balance a single day of previous behaviour. The second, the difference between R_0_ on lockdown and 1 is the proportion that contact under lockdown could be increased without causing a resurgence of the epidemic. As the continuing behaviour is quite different from that prevented, partly because much of what is permitted is within the domestic environment and most of that restricted is external to it, this measure is relatively uninformative and not discussed further. The third measure was calculated as:

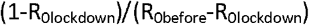

Provided R_0lockdown_ is less than both 1 and R_0before_, this gives an estimate of the proportion of the behaviour, prevented by the lockdown, that can be resumed and result in an overall R_0_ equal to 1. The first and third approaches give different results because daily changes combine multiplicatively. An example that demonstrates this would be for a disease with a generation time of 1 week that had R_0_ of 2 initially and ½ under lockdown: alternate weeks of doubling and halving would oscillate around a constant value, with a mean contact rate of 1.25, higher than the continuous R of 1 that would produce stability. Counterintuitively, that suggests more activity overall might be possible under a strategy of intermittent lockdown. Confidence intervals around each estimate were generated by drawing 1000 values from the relevant model parameter distributions.

Results are presented for 73 countries. Because so many countries were considered, some results can be expected to appear significant by chance and caution is needed in interpreting individual results. The discussion below will therefore concentrate on general patterns across multiple countries.

## Results

Figure 1 shows trajectories for the five countries with the highest number of deaths in this dataset, and table 1 parameter estimates for them. The graphs for the remaining countries are in the supplementary material. It can be seen that the chosen intervals of exponential growth are in the early stages of the epidemic, which fits with the behavioural change to be expected, and that was intended, as regulations were imposed and public awareness of the urgency of the problem spread.

**Figure 1:**
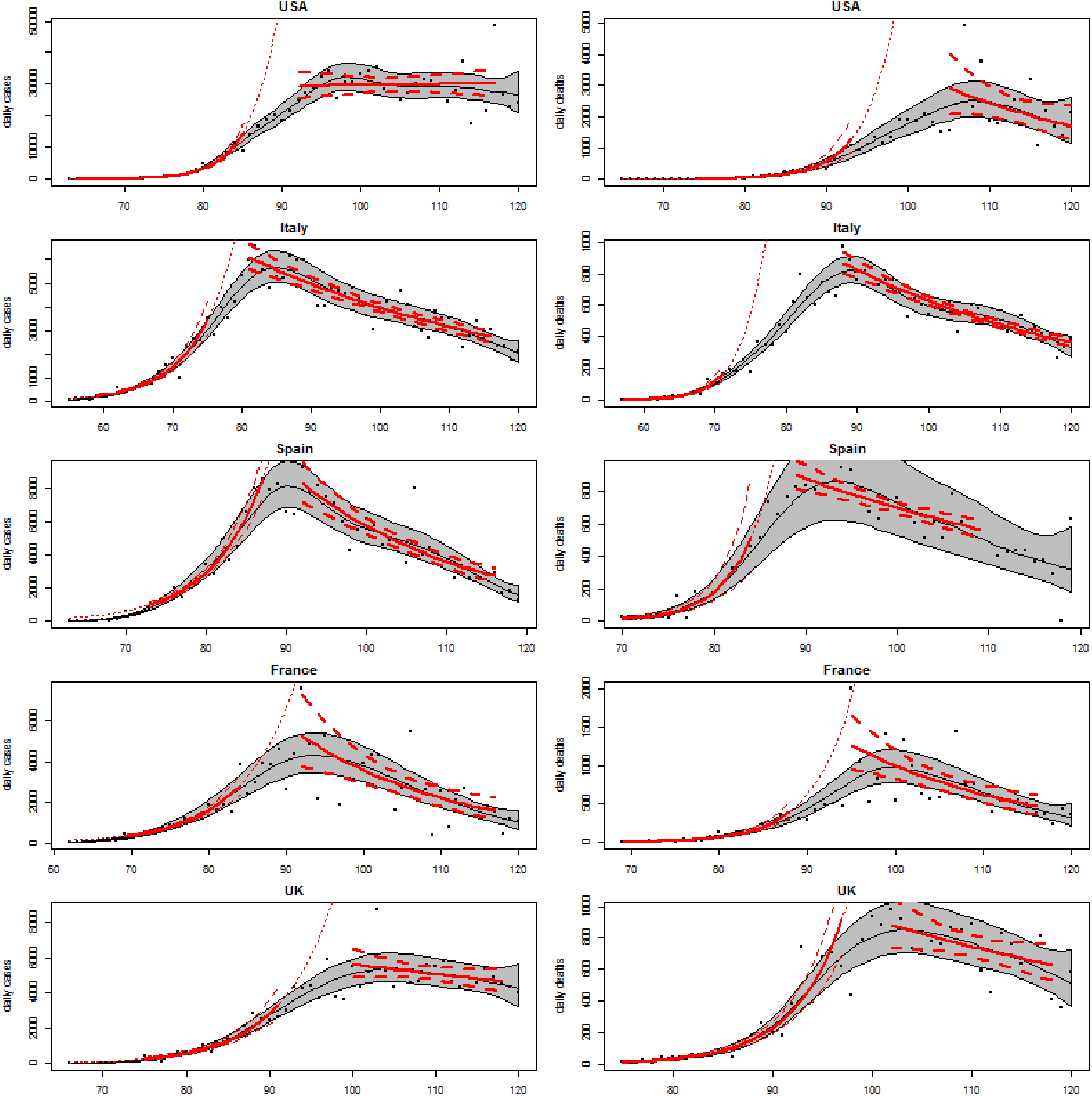
Trajectories of cases of, and deaths from, COVID-19.

**Table 1:** R_0_ estimated in the models of increasing and declining phases based on mortality data and confimed cases of COVID-19 19 up to 29th April2020.

| Country | Confirmed cases |  |  | Deaths |  |  |
| --- | --- | --- | --- | --- | --- | --- |
|  | N | Before | After | N | Before | After |
| USA | 1012583 | 3.6 (3.4, 3.8) | 1.01 (0.96, 1.06) | 58355 | 3.2 (2.7, 3.7) | 0.83 (0.67, 1.01) |
| Italy | 201505 | 2.2 (2.0, 2.4) | 0.89 (0.87, 0.91) | 27359 | 3.7 (3.1, 4.4) | 0.87 (0.85, 0.89) |
| France | 126835 | 2.0 (1.9, 2.1) | 0.77 (0.68, 0.87) | 23660 | 2.7 (2.4, 3.0) | 0.78 (0.68, 0.88) |
| Spain | 212203 | 2.0 (1.8, 2.2) | 0.78 (0.73, 0.83) | 23412 | 3.2 (2.4, 4.1) | 0.88 (0.85, 0.93) |
| UK | 161145 | 2.1, (1.8, 2.3) | 0.95 (0.88, 1.01) | 21678 | 2.5 (2.2, 2.7) | 0.90 (0.81, 0.99) |

For cases in the USA, the second exponential phase is estimated to probably be another period of exponential growth, though at a slower rate than the initial period. However, daily mortality does seem to have declined in the USA. The confidence intervals around the exponential models are noticeably narrower than those around the GAMs, this is largely due to the information the GAMs require for the extra parameters that describe their curvature.

Visual inspection of the plots suggest that the method has selected subperiods that do not seem representative for cases in 5 countries (China, Greece, Japan, Morocco, and Turkey) and for mortality for 3 (Indonesia, Japan, and Turkey). These results are included, but indicated, in the figures. The models ignored the first, small, outbreak in Singapore and picked up only the larger growth since then.

For Armenia, Belarus, Brazil, Colombia, Kuwait, Mexico and Panama, the initial exponential growth in numbers of cases appears to have been followed by a brief period of decline before the growth resumed. This pattern also appears in the mortality data for Brazil, India and Poland. And the models miss an early period of rapid decline in new cases in Denmark, focussing on the subsequent slower decline. Conversely, for cases in the Netherlands, and mortality in Germany, an initial period of slow decline was selected despite the later data appearing to decline more rapidly. However, while interesting, all these patterns should be treated with caution.

Supplementary Table 1 contains the estimated slopes, and standard errors, for the two best exponential models of cases and mortality trajectories in each model. As doubling times are more immediately interpretable, these are shown (Figure 2A). Many of these are too imprecise to be useful. However, for those countries with sufficient data, estimates of doubling times from mortality data are generally around 2 to 5 days. The estimates of halving time under lockdown (Figure 2B) are generally over 7 days, and much higher than the equivalent pre-lockdown doubling times. This impression is confirmed by inspection of individual trajectories, most of which decline more slowly than they increase.

**Fig 2:**
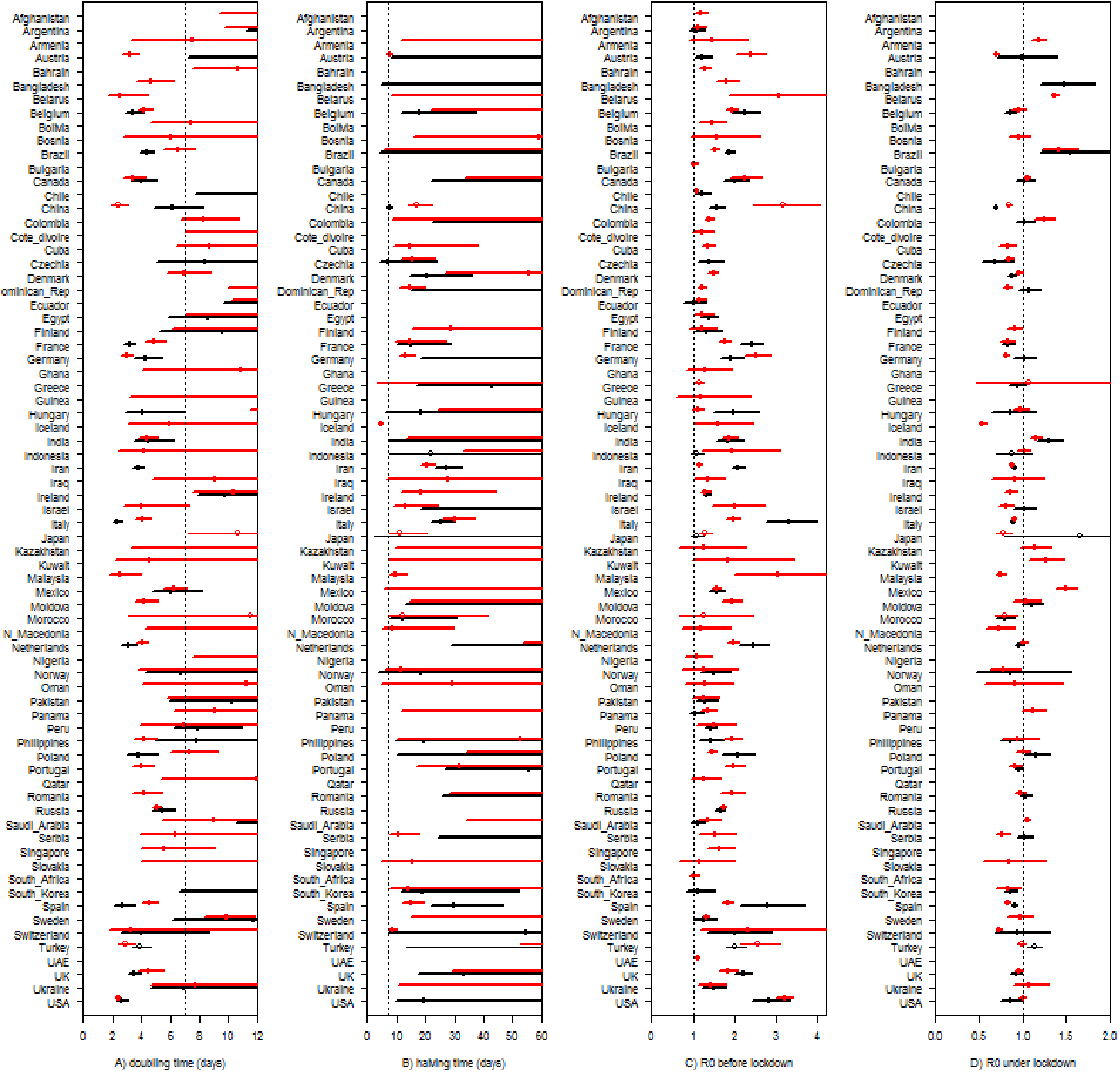
Doubling times and R0 for exponential phases of outbreaks of COVID-19.

The dots are numbers of new cases (left) or deaths (right) reported to the ECDC for each day up to 29^th^ April 2020. Each grey pipe shows the 95% confidence interval around a smooth trajectory (black line) estimated by a generalised additive model. In red are increasing and decreasing exponential patterns (mean & 95% CI) fitted to subsets of the data. Details of the models are in the main text.

Black points are estimates based on mortality data, red are based on confirmed cases. The lines are 95% confidence intervals. Thin lines and hollow points indicate countries where plots of the modelled trajectories led to subjective doubts of the model fits. Gaps occur where either exponential phase was identified. Supplementary Table 1 contains all these data. A) doubling time in the first (pre lockdown) exponential phase of the epidemic. Lines that meet 0, the left hand side of the box, indicate a more than 2.5% chance that the epidemic was slowing over this period. B) halving time in the second (locked down) exponential phase. The vertical dotted line is at 7 days in panes A and B, and shows that almost all declines were slower than the preceding increases. C) The basic reproduction number, R0, for COVID-19 in each country during the first exponential phase. D) R0 under lockdown. The vertical dotted line at 1 is a guide to highlight how little evidence there is for lockdowns having reduced R0 below this number.

The estimates of R_0_ contain essentially the same information as those of doubling times. The definition of the initial exponential period requires all the point estimates of R_0_ to be greater than 1, though the lower bounds of some of the confidence intervals fall below that threshold (Figure 2C and Supplementary Table 1). Almost all the values under lockdown (Figure 2D) are between 0.6 and 1.5. There are only 6 countries (Italy, France, Iran, China, Czechia, and Morocco) where the upper bound of the estimate of R_0_ for the mortality data is below 0.9, though this is also true for the case data of 15 countries.

Because it is such a short time since most countries have introduced restrictions and changed the behaviour of their populations, there is a lot of uncertainty in the estimated trajectories under lockdown. But what evidence there is (Figure 3) suggests that periodical release of lockdown beyond one week in each month is very likely to result in the acceleration of the epidemic in many countries. And even this is based on the assumption that behaviour during intermittent releases from lockdown resembling typical behaviour in the period before the behavioural changes associated with the COVID19 epidemics.

**Fig 3:**
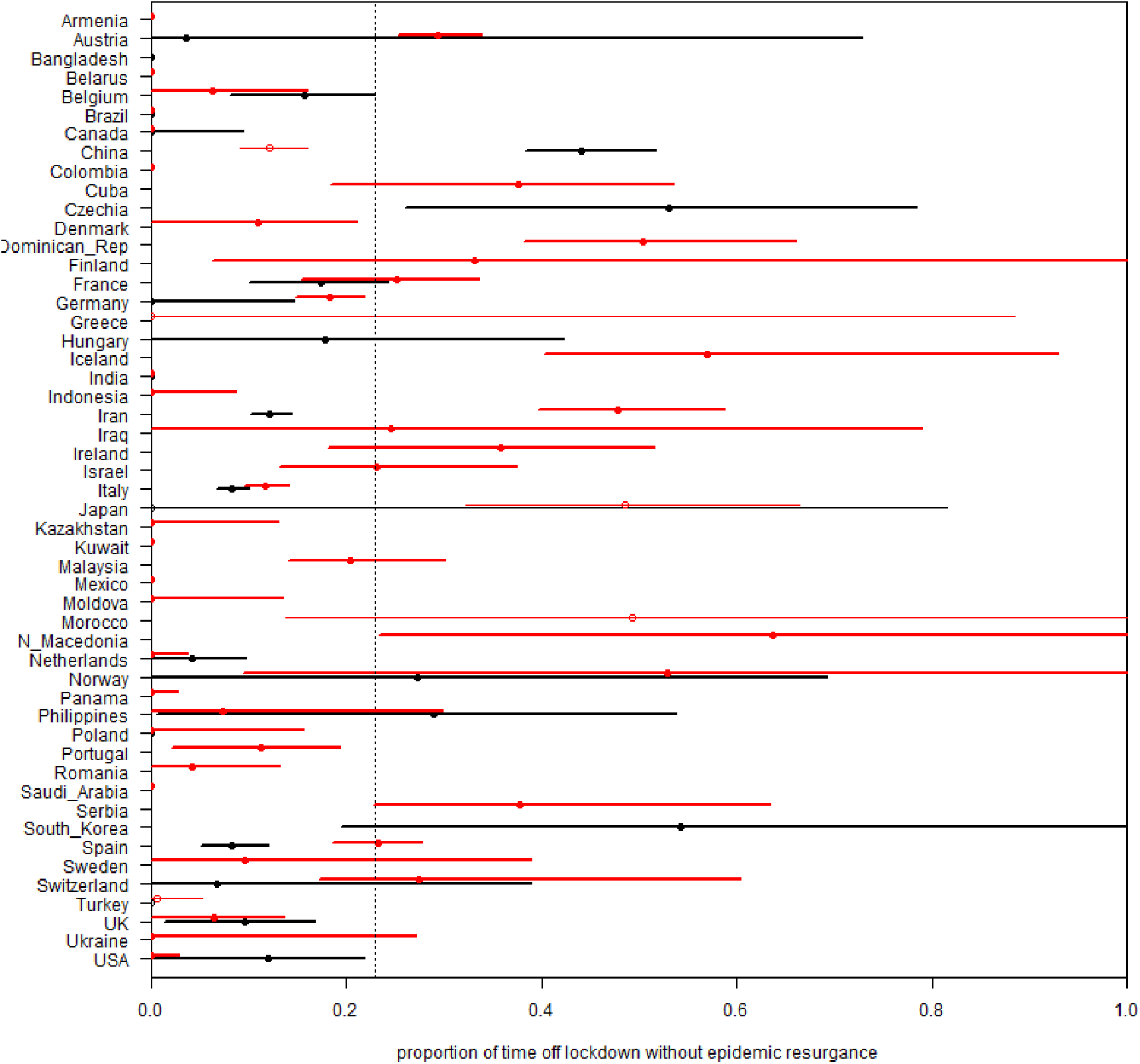
Estimates of the proportion of time that countries could have in their pre-lockdown state without the spread of COVID-19 resuming.

For each country the exponential rate calculated before lockdown was divided by that under lockdown to give a mean and 95% confidence interval. Black is again based on mortality and red on confirmed cases. Thin lines and hollow points indicate countries where plots of the modelled trajectories led to subjective doubts of the model fits. Lines that meet 0, the left hand side of the box, indicate a more than 2.5% chance that the epidemic was actually spreading faster over the second period than the first. The vertical dotted line shows that there is little evidence for believing that relaxing lockdown for one week (during which previous behaviour would be resumed) each month would be consistent with stopping the epidemic.

Similarly, these data show that few countries seem to be in a position to confidently ease their lockdowns, (Figure 4).Only for Italy, France, Spain, Iran and China, do models of both the case and mortality data exclude 0 from the 95% confidence intervals around their estimates of the proportion of easing of restrictions that would not raise R_0_ above 1. For five other countries, the mortality models capture their reported success in containing COVID-19, but the models of numbers of cases do not. Resuming even 20% of currently prevented behaviour looks extremely ambitious in most countries, with even a 5% easing appearing potentially risky in many of them.

**Fig 4:**
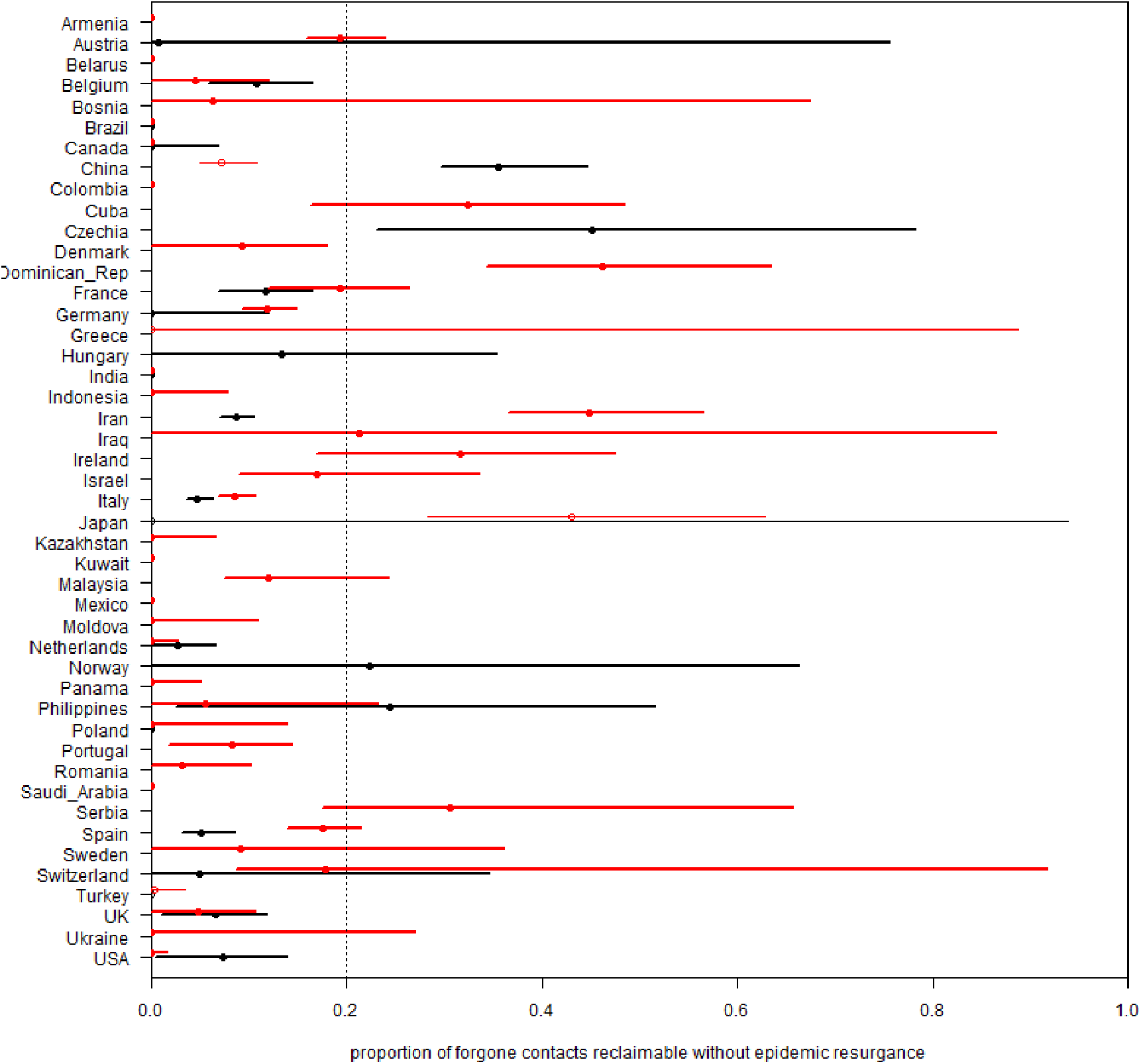
The scope for partial easing of lockdown while containing the spread of COVID-19.

One minus R_0_ under lockdown divided by the difference between the R_0_’s before and under lockdown is an estimate of the proportion of the behaviour, that lockdown has prevented, that can be resumed without increasing R_0_ past 1, and restarting epidemic spread. Each row is the mean and 95% confidence interval for a country, black uses mortality data, red confirmed cases. Thin lines and hollow points indicate countries where plots of the modelled trajectories led to subjective doubts of the model fits. Lines that cut the left hand edge (0) indicate countries for whom lockdown may not have achieved stability. The vertical dotted line at 0.2 is a guide to highlight that there is little evidence for it being sustainable to resume more than 20% of discontinued activity without the epidemic resuming its spread.

## Discusssion

These analyses are, by their nature, provisional and attempting to provide estimates and predictions from limited datasets. The models differ from most of the infectious disease models that have been applied to the COVID epidemics: they are based entirely on behaviour observed during the present pandemic of SARS-CoV-2 and do not incorporate any assumptions based on other respiratory viruses. Only the conversion of the slope parameters to estimates of R_0_ uses a previously estimated distribution of generation times. This simplicity avoids reliance on the uncertain assumptions, necessary for more traditional epidemiological models, instead increasing the confidence intervals are the estimates. Despite this, a surprisingly clear picture is visible: if COVID-19 had been only slightly more transmissible the current lockdown measures would have been unable to halt the epidemic in Europe. It is also not clear that it actually has been stopped in many data-rich countries. Where it has been stopped, the margins for loosening the controls are frighteningly thin. And the slow rates of decline in mortality suggest uncomfortable lower bounds on future mortality: sustaining a 10% per day decline, a rate faster than the best estimate for almost all the lasting declines in these data, implies that there will be a total of another 9 times as many deaths to come as were reported yesterday; 5% per day increases that to 19 times.

There has been talk of lockdown being “a cure worse than the disease”, but it is not a cure. At 5% per day it will take 35 days to claw back from 600 to 100 deaths per day; and another 45 to then get down to 10. If 10% per day could be sustained it would only require 18 plus 20 days. But many countries are not yet clearly past, or even close to, the peak. That suggests easing in the near future will imply continuing mortality, and substantial easing is very likely to require the rapid re-imposition of lockdown.

R_0_, or more precisely R_t_, seems to be the key to this problem: until and unless a vaccine or effective treatment becomes available, we need a liveable way to keep R below 1. Lockdown, to a point beyond what most societies would have previously imagined accepting, can barely contain the disease’s spread. It is hard to see it continuing indefinitely. These data suggest that unless a vaccine becomes rapidly available, discussions around exit strategy from current restrictions therefore need to move on from optimistic concepts of returning rapidly to normal activity. The data is more consistent with a need to adopting a “new normal” that can provide the optimal balance between allowing economic activity while ensuring very substantial reductions in prior social contacts (at least 90% reductions according to our best estimates). It is beyond the scope of this paper to describe what the components of a new normal may be but discussions will include continuing social distancing, public use of face-coverings, testing, tracking and isolating infected individuals and contacts and widespread screening of asymptomatic individuals among other considerations.^13–15^

In summary, a simple analysis based on the behaviour of the SARS-CoV-2 pandemic to date across 73 countries suggests remarkably consistent effects of both exponential growth and slow decline in cases and mortality. Without a vaccine, these estimates are incompatible with a return to previous activities post “lockdown”.

## Data Availability

The data was obtained from the ECDC, and is publically avaliable at: https://www.ecdc.europa.eu/en/publications-data/download-todays-data-geographic-distribution-covid-19-cases-worldwide

## Supporting Information

1. Supplementary Table 1: Parameter estimates from models of exponential trajectories within COVID-19 outbreaks for 73 countries, though most have very limited or imprecise data
2. Supplementary plots of trajectories, similar to those in Figure 1, for all the countries listed in Supplementary table 1
3. The R code used for these calculations.

